# RISKS ASSESSMENT OF CARDIOVASCULAR DISEASES AMONG HIV/AIDS PATIENTS RECEIVING ANTIRETROVIRAL THERAPY AT UNIVERSITY TEACHING HOSPITAL OF BUTARE CHUB

**DOI:** 10.1101/2024.12.27.24319701

**Authors:** Nshimiyimana Charles, Nemeyimana Patrick, Uwumuremyi Fabrice, Karenzi Valens, Bakunzibake Pierre, Mugisha Emmy

**Affiliations:** University of Rwanda-center of excellence in biomedical engineering and ehealth; Ines Ruhengeri; University of Rwanda-college of medicine and health sciences

**Keywords:** HIV/AIDS, cardiovascular diseases, antiretroviral therapy, lipid profile, CHUB, Rwanda, cross-sectional study, hypertriglyceridemia, HDL cholesterol, LDL cholesterol

## Abstract

This study assessed the risks of cardiovascular diseases among HIV/AIDS patients receiving antiretroviral therapy (ART) at the University Teaching Hospital of Butare (CHUB) in southern Rwanda. A cross-sectional design was employed from January to February 2021. A sample of 199 participants was systematically selected, including HIV-positive patients on ART and a control group. Blood samples were analyzed for lipid profiles using standard spectrophotometric procedures. The study adhered to strict ethical considerations, including informed consent and participant confidentiality. Data analysis was performed using SPSS, with a p-value of ≤0.05 considered statistically significant. Results indicated demographic disparities, with 66% female participants and 72% on first-line ART. Triglyceride levels showed 42% within normal ranges, 26% borderline high, and 32% hypertriglyceridemia. For high-density lipoprotein cholesterol (HDL-C), 61.3% were normal, 19.3% classified as low-risk, and 19.3% had hypercholesterolemia. Low-density lipoprotein cholesterol (LDL-C) results revealed 87.6% within normal ranges, 7.3% borderline high, and 8% with hypercholesterolemia. Total cholesterol analysis is ongoing. Ethical approval was obtained, though the study faced challenges such as a limited sample size and movement restrictions due to COVID-19. Despite limitations, the findings highlight the importance of monitoring lipid profiles in HIV/AIDS patients on ART to reduce cardiovascular risks and inform clinical interventions.

## CHAPTER I: INTRODUCTION

### 1.1 DEFINITION OF KEY TERMS

#### Lipid profile

A pattern of lipid in the blood. It includes level of TC, HDL-c, TG and LDL-c (Feeneyand Mallon, 2011).

#### Cholesterol (also known as total cholesterol)

A fatlike substance that is a building block of the outer layer of cells (cell membranes). It is essential to the establishment of bile acids, cell membranes, vitamin D and certain hormones. Cholesterol is not dissolved in the blood, but is transported in the bloodstream as water-soluble molecules known as lipoproteins. The lipoproteins are characterized by their density: high density lipoprotein (HDL) and low-density lipoprotein (LDL) (Tadewos *et al*., 2012).

Hypercholesterolemia: high amount of cholesterol in the blood (Bekolo *et al*., 2014).

#### High density lipoprotein (HDL)

A fat-like substance that transports cholesterol from the tissues of the body to the liver so it can be excreted in the bile. HDL is the so-called “good cholesterol”; the higher the HDL cholesterol level, the lower the risk of coronary heart disease (CHD)(Feeney and Mallon, 2011).

#### Low density lipoprotein (LDL)

A fat-like substance that transports cholesterol from the liver to the tissues of the body. LDL is the so-called “bad” cholesterol; elevated LDL levels are associated with increased risk of coronary heart disease (CHD) (Mbuagbaw *et al*., 2010).

#### Risk

Something that increases a person’s chances of developing a disease

#### Triglyceride

The major form of fat. A triglyceride consists of three molecules of fatty acid combined with a molecule of the alcohol glycerol. Triglycerides serve as the backbone of many types of lipids (fats). Triglycerides come from foods and are also produced by the body. Triglyceride levels do not provide clinically significant information about the risk of coronary heart disease (CHD) beyond that provided by levels of HDL and LDL cholesterol (Tadewos *et al*., 2012).

#### Hypertriglyceridemia

High level of triglyceride in the blood (Nsagha, Assob, *et al*., 2015).

#### Cardiovascular diseases

Refer to the heart and blood vessels diseases (Grinspoon and Carr, 2005).

#### Atherosclerosis

Is a condition where the arteries become narrowed and hardened due to the build-up of plaque around the artery wall (Kramer *et al*., 2008).

#### Antiretroviral therapy (ART)

Mixture antiretroviral regimens that hostilely decrease HIV viral multiplication to halt the progress of HIV disease. The usual ART regimen combines three or more different drugs. These treatment regimens can reduce the amount of virus so that it becomes undetectable in a patient’s blood (Mbuagbaw *et al*., 2010).

### 1.2 BACKGROUND

HIV/AIDS remains one of the major health problems in the most parts of the world with increased morbidity and mortality as major public health complications in both developed and developing countries. Worldwide around 35.3 million people are living with HIV and in 2012, 22.9 million are living in Sub Saharan Africa. It is predictable that greater than 14000 people are infected daily with HIV and other 11000 are dying daily due to human immunodeficiency virus (WHO,2005).The estimated number of AIDS related deaths in 2017 was predictable to be 1.6 million with adults being 1.4 million (Alberts *et al*., 2005).

It has been mentioned that 600,000 among Cameroonian people were predictable to be living with HIV and among them 46.7% or 280,000 were suitable for ART for a better future lifesaving jointly,1.2million people living with HIV/AIDS has been mentioned to be in Ethiopia (Tadewos *et al*., 2012).

HIV epidemic in Rwanda is generalized, with prevalence of 3.2% in adult population aged from15 years up to 45 years and it is commonly highest among women aged at 40-44(8%) and men age 45-49(9%). The higher prevalence of HIV/AIDS is present mostly in women than men and present highly in urban than rural areas (Rwanda national strategic plan on HIV and AIDS 2009-2012). It has shown that Kigali city has higher prevalence of 7.3% meanwhile the lowest prevalence of HIV in Rwanda is found in Eastern province at 2.1% (Biraguma, Mutimura and Frantz, 2018).

As reported by WHO in 2018, approximately 9.7 million people in low- and middle-income countries were receiving antiretroviral therapy by June 2011 as response of HIV/AIDS reduction and proper life handling(Feeney and Mallon, 2011). However, several reports have documented increased risk of cardiovascular diseases (CVD) such as obesity, abnormal lipid profile like hypertriglyceridemia, high low-density lipoprotein (LDL-c) and low high-density lipoprotein cholesterol (HDL-c)) in HIV individuals under HAART-treated regime (Calza, Manfredi, Pocaterra, & Chiodo, 2008; Nsagha *et al*., 2015; Souza, Luzia, Santos, & Rondó, 2013). The prevalence of cardiovascular diseases in PLWHA from published studies may varies from region to regions (Grinspoon and Carr, 2005).

Treatment regimens including nevirapine, norvir, abacavir, lamivudine and efavirenz have also been reported to induce lipid derangements and increase the risks of CVD. ART can induce raised levels of total cholesterol (TC), LDL-cholesterol (LDL-c) and triglycerides (TG), and variables effects on HDL-cholesterol (HDL-c) levels (Karangwa *et al*.,2006). Also, several reports have documented increased prevalence of hypertriglyceridemia, and low HDL-c in HIV individuals receiving ART (Alberts *et al*., 2005; Souza *et al*., 2013).Side effects and toxicities are associated with these highly effective therapies and there is rising alarm that the metabolic complications such as liver, kidney and heart are associated with HIV and antiretroviral therapy which may lead to an increased risk for cardiovascular diseases(Mbuagbaw *et al*., 2010).

Highly active antiretroviral therapy (HAART) regimen has profoundly decreased morbidity and mortality rates associated with HIV infection despite the reported increased risk of cardiovascular disease (CVD(Oka *et al*., 2012). With the reference to World Health Organization (WHO), PWHA under HAART are at high risk to develop lipid profile metabolic disorders, thus recommended that further studies related to this association. A combination of zidovudine(AZT), lamivudine(3TC), efavirenz (EFV) and Nevirapine (NVP) as the first line regimes, abacavir (ABC), lipinovir (LPV), atozovir (ATV) and tenofovile (TDF) as second line regimens and (Raltegravir) RAL+(Darunavir)DRV+ (Ritonavir)RTV for third line used in Rwanda in 2007 and it was revised in 2019 and zidovudine was replaced by tenofovile (TDF) (MoH,2007) and today in Rwanda these medications are being used nevirapine, norvir, abacavir, and lamivudine. Therefore, the current study aims to the risk assessment of cardiovascular diseases among HIV infected individuals receiving highly active antiretroviral therapy regimen at University Teaching Hospital of Butare (CHUB).

### 1.3. PROBLEM STATEMENT

HIV/AIDS is a major health problem in the Sub-Saharan Africa, with increased morbidity and mortality ranging from 24.2 to 44% (Fortes D et al,2011). In Rwanda it comes to be among the endemic disease with prevalence of 3.1% (Kayirangwa *et al*., 2006). With the use of ART, this mortality rates decreasing with an increases the risk factors of CVD (Alberts *et al*., 2005).

The routine treatment of HIV infection includes nevirapine, norvir, abacavir, lamivudine and efavirenz which play role in inducing lipid profile derangements one of the major risks of CVD. In Rwanda HIV/AIDS receiving HAART are 97.5% and can persuade raised levels of total cholesterol (TC), LDL-cholesterol (LDL-c) and triglycerides (TG), and variables effects on HDL-cholesterol (HDL-c) which may contribute to the development of cardiovascular diseases (Bekolo *et al*., 2014).

Based on the recent studies conducted, it estimated that HIV/AIDS in African receiving ART are subjected to increased level of cholesterol but within their study they did not show any relationship between this increased lipoprotein levels with risk of cardiovascular diseases (Grinspoon and Carr, 2005). In Rwanda, the risk of cardiovascular diseases among PLWHA is not well known. Therefore, this study aims to determine the lipid profile and their association with ART used in the treatment and control HIV in people living with HIV (Tadewos *et al*., 2012).

### 1.4. RESEARCH QUESTION

How are the lipid profile and its association with HAART regimens in HIV infected individual receiving antiretroviral therapy at university teaching hospital of Butare (CHUB)?

### 1.5. OBJECTIVE OF THE STUDY

#### 1.5.1. MAIN OBJECTIVE

Risks assessment of cardiovascular diseases among HIV/aids patients receiving antiretroviral therapy at University Teaching Hospital of Butare (CHUB)

#### 1.5.2. SPECIFIC OBJECTIVES OF THE STUDY

1. To determine the lipid profile -: total cholesterol (TC), LDL-cholesterol (LDL-c) and triglycerides (TG), and HDL-cholesterol (HDL-c) levels as biochemical risks markers of CVD in HIV/AIDS receiving ART at CHUB.
2. To determine the association between lipid profile and ART treatment regimens used in HIV/AIDS patients.

### 1.6. SIGNFICANCE OF THE STUDY/JUSTIFICATION

The information which will be generated from the study will support the Ministry of Health as well as different referral hospitals and district hospitals in policies development regarding the prognosis and management of cardiovascular diseases in HIV/AIDS patients receiving antiretroviral therapy. Therefore, detection of lipid profile in PLWH may be added to the routine biological assessment to determine the risks of CVD for early intervention and prevention of the disease.

Failure to appreciate possible emerging problems of CVDs in PLWHA in SSA may mitigate the gains from antiretroviral therapy in African region with a high disease burden (Grinspoon, S. and Carr, A. 2005). Determination of cardiovascular diseases and ART treatment regimens like combination of zidovudine(AZT), lamivudine(3TC) and Nevirapine (NVP) association and appropriate intervention strategies where necessary as required and would allow for assessment of the impact of HAART on CVD risk (Nsagha *et al*., 2015).

## CHAPTER II: LITERATURE RIVIEW

### 2.1 INTRODUCTION

Cardiovascular disease (CVD) is a major cause of morbidity and mortality worldwide, with the lifetime risk exceeding 60%.1 More than 2200 Americans die of CVD daily, 1 death every 40 seconds (Friis-Møller N, 2003). A third of CVD deaths occur before 75 years of age, which is less than the average life expectancy of 78.8 years.1 Thus, prevention of CVD is a public health priority.

Through respects to WHO updated heart diseases management in 2006 stated that CVDs are the number one cause death globally where more people die annually from CVDs than from any cause(Glass *et al*., 2006). An estimated 17.9 million people died from CVDs in 2016 representing 31%of all global deaths of these, 85% are due to heart attack and stroke. Over three quarters of CVDs death take place in low- and middle-income countries (Hemkens & Bucher, 2014). Out of the 17 million premature deaths (under the age of 70) due to non-communicable diseases in 2015,82% are in low- and middle-income countries in which 37% are due to CVDs definitely untreated cardiovascular diseases mostly due to ART drug consumption are silently killing most of the users in which might be confused as it is the HIV/AIDS that leading to the loss of life(Bloomfield *et al*., 2011).

With refer to the study conducted in South Africa, there was a high prevalence of High-density lipoprotein-cholesterol was relatively high, whereas 42.3% of women and 28.5% of men had low density lipoprotein-cholesterol levels of 3mmol/l or more (Zieman, 2007). Hypertension (blood pressure 140/90mmHg) was found in 25.5% of women and 21.6% of men. According to the Framingham formulae, 18.9% of women and 32.1% of men had a 20% or higher chance of having a CVD. Therefore, there was a high prevalence of chronic heart disease and risk factors in the rural, poor black community in Limpopo, South Africa. Consequently, the population had a higher than expected risk of developing a CVD event in within a short period of time when compared with similar studies in black Africans(Risk & Report, 2015).

Within a study steered at Cameroon, HIV infection was most prevalent in subjects aged 31 to 49 years. Most of the HIV-positive patients belonged to Centres for Disease Control and Prevention categories B (43.0%) and C (30.23%) (Bekolo *et al*., 2014). Compared with control subjects, patients with CD4(Cluster of Differentiation) counts of 50cells/mL had significantly lower TC (Po0.0001) and LDL-C (Po0.0001) but significantly higher triglyceride (TG) values (Po0.001) and high HDLC/LDLC (P50.02); patients with CD4 counts of 50–199cells/mL had significantly lower TC (Po0.001) and significantly higher TG values (Po0.001); patients with CD4 counts of 200–350cells/mL had significantly higher TG (P50.003) and a higher atherogenicity index for TC/HDLC (Po0.0002) and HDLC/LDLC (P50.04); and those with CD4 counts 4350 cells/mL had a higher atherogenicity index for TC/HDLC (Po0.0001) and HDLC/LDLC (Po0.001) (Bekolo *et al*., 2014).

HDLC was significantly lower in HIV-positive patients irrespective of the CD4 cell count. Thus, HIV infection is associated with dyslipidaemia later to CVD in patients receiving ART, and becomes increasingly debilitating as immunodeficiency progresses. HDL-C was found to be lower than in controls in the early stages of HIV infection, while TG and the atherogenicity index increased and TC and LDLC decreased in the advanced stages of immunodeficiency (Grinsztejn *et al*., 2013).

Upon to study conducted by MUTIMURA, where A HAART regimen of stavudine, lamivudine, and nevirapine was used by 81.6% of subjects; none received protease inhibitors. Dyslipidaemia was observed in 34% (48.5% in urban groups and 17.3% in rural groups) of subjects, with a prevalence of 69.6% in those receiving HAART for >72 weeks. Peripheral lipoatrophy combined with abdominal dyslipidaemia was observed in 72% of l dyslipidaemia subjects (Biraguma *et al*., 2018).

HIV-positive adults with lipodystrophy had a significantly higher value-to-hip ratio (WVR; 0.99 ± 0.05 vs. 0.84 ± 0.03: P < 0.0005) than HIV positive non-dyslipidaemia adults. Total cholesterol concentrations (median in mmol/L) were significantly higher in the HIV-positive adults with dyslipidaemia (3.60 [1.38]) than in HIV-positive non-dyslipidaemia adults (3.19 [0.65]; P < 0.005) and control (3.13 [0.70]; P < 0.0005) groups(Feeney & Mallon, 2011). Impaired fasting glucose was observed in 18% of HIV-positive adults with dyslipidaemia, 16% of HIV-positive non-dyslipidaemia adults, and 2% of controls, but insulin levels did not differ. finally, African subjects with dyslipidaemia have increased glucose, and cholesterol levels (Nsagha, *et al*., 2015).

### 2.2. DYSLIPIDAEMIA

CVD risk factors occur more frequently in combination than in isolation and have been associated with concurrent dyslipidaemia. There is mark that dyslipidaemia act synergistically to increase CVD risk. Based on cholesterol treatment guideline recommendations, clinicians should evaluate a patient‟s overall CVD risk when considering cholesterol lowering therapy because many patients with hypertension, but without elevated LDL-C, may benefit from statin therapy(Thomas F. Lüscher, 2018). Accordingly, a comprehensive approach to CVD risk factor modification, especially for hypertension and dyslipidaemia, is essential to maximize the reduction in CVD. Although some CVD medications have modifying effects on blood pressure and cholesterol, these effects are generally small and overshadowed by the reductions in CVD events. Moreover, novel lipid lowering therapies like PCSK-9 inhibitors show even greater reductions in lipids without adverse changes in blood pressure, effects that may translate into further reductions in CVD among selected hypertensive patients with suboptimal lipid levels (John W,2018)

Dyslipidaemia is a prerequisite to the development of CVD, Only in populations with lifelong with high level of cholesterol in the blood(Bloomfield *et al*., 2011). As with blood pressure, lipid levels track through childhood. Autopsy studies have consistently demonstrated the adolescent onset of the atherosclerotic process, and the Bogalusa Heart Study has confirmed the strong association between antemortem cholesterol levels and post-mortem atherosclerosis in adolescents and young adults (Elaine M,2018) definitely, dyslipidaemia has been noted as one of the major contributing factors of cardiovascular diseases.

### 2.3. DYSLIPIDAEMIA AMONG HIV POSITIVE PATIENTS

Increasing incidence of dyslipidaemia has been recognized among HIV infected individuals. HIV infection per se is associated with lipid alteration including total cholesterol and it components (low HDL- and LDL-cholesterol and hypertriglyceridemia)(Oka *et al*., 2012). The use of HAART has been associated with elevation of serum lipids above that of its pre-treated levels, except for HDL cholesterol. The prevalence of dyslipidaemia in HIV infected patients ranges from 28% to 80% and includes hypotriglyceridaemia (40-80%), hypercholesterolemia (10-50%) and mixed forms Insulin resistance has been considered the link between dyslipidaemia and other metabolic disturbances within metabolic syndrome such as abdominal obesity, diabetes, heart diseases, and hypertension(Thomas F. Lüscher, 2018).

Clinically, HIV infection and its treatment are associated with abnormalities in lipid metabolism. Before the availability of highly active antiretroviral therapy (Grunfeld *et al* 2009). found that patients with AIDS had elevated plasma triglyceride and free fatty acid levels whereas HIV-infected patients without AIDS had decreased total cholesterol and HDL-cholesterol (Sniderman *et al*., 2011).

In the Multicenter AIDS Cohort Study (MACS) cohort, total cholesterol, HDL-cholesterol, and LDL-cholesterol levels declined following HIV seroconversion. In these patients, total cholesterol and LDL-cholesterol levels rose but HDL-cholesterol remained decreased after initiation of highly active antiretroviral therapy (Hadigan *et al*., 2001).

In the Swiss HIV Cohort Study, HIV protease inhibitor use was found to be associated with increases in plasma total cholesterol and triglycerides. Discontinuation of antiretroviral therapy in the study resulted in a decline in total cholesterol and LDL-cholesterol, but HDL-cholesterol declined as well, leading to an unfavorable increase in the total/HDL-cholesterol ratio (Mbuagbaw *et al*., 2010).

Recently, some evidences showed that cardiovascular risk in HIV patients was associated with lower levels of small and large HDL-particle concentration independently of other cardiovascular risk factors. Patients with HIV-associated fat redistribution are even more likely to have dyslipidemia, possibly related to excess visceral adipose tissue accumulation. Among patients with HIV-associated lipodystrophy, found the LDL mean particle size to be less than 20.5 nm and therefore exhibiting a proatherogenic pattern (Grinsztejn *et al*., 2013).

Dyslipidemia in HIV patients may contribute to their increased risk of cardiovascular disease. The identification and treatment of lipid abnormalities are important in HIV patients as they are modifiable cardiovascular risk factors (Sniderman *et al*., 2011).

### 2.4 ATHEROSCLEROSIS

Atherosclerosis is the narrowing of arteries due to plaque build-up on the artery walls. Arteries carry blood from the heart to the rest of the body (Triant *et al*., 2007). They are lined with a thin layer of cells that keeps them smooth and allows blood to flow easily. This is called the endothelium. Atherosclerosis starts when the endothelium becomes damaged, allowing the harmful type lipid panel to build up in the artery wall(Bloomfield *et al*., 2011).

The body sends a type of white blood cell to clean up this cholesterol, but, sometimes, the cells get stuck at the affected site. Over time, plaque can build up, made of cholesterol, macrophages, and other substances from the blood (Germany *et al*., 2012). Sometimes, the plaque grows to a certain size and stops growing, causing the individual no problems. However, sometimes, the plaque clogs up the artery, disrupting the flow of blood around the body. This makes blood clots more likely, which can result in life-threatening conditions.

In some cases, the plaque eventually, breaks open. If this happens, platelets gather in the affected area and can stick together, forming blood clots. This can block the artery, leading to life-threatening complications, such stroke and attack. The condition can affect the entire artery tree, but mainly affects the larger, high-pressure arteries (Kiyotaka Hao, 2012).

### 2.5. HIV INFECTION INCREASES ATHEROSCLEROSIS-ASSOCIATED CVD IN THE ART PATIENTS

The use of ART has increased the survival of HIV^+^ patients to close to that of the general population. Although fewer HIV patients are dying of AIDS-related complications, the prevalence of non-AIDS-related comorbidities, including atherosclerosis-associated CVD(Thomas F. Lüscher, 2018), remains increased compared with HIV^−^ controls. CVD is the second leading cause of non-AIDS-related mortality in the United States and third in Europe among HIV^+^ patients. Data suggest that HIV^+^ patients have a higher prevalence of traditional risk factors, including hypertension, diabetes, and dyslipidaemia. There is extensive evidence to suggest that even when controlled for these traditional cardiovascular risk factors, HIV^+^ patients are still at a higher (1.5- to 3-fold) risk of developing CVD (Triant *et al*., 2007).

Two landmark clinical trials have provided valuable information in regard to ART and its timing of ART in HIV infection as they relate to CVD: the SMART (Strategies for Management of Antiretroviral Therapy) and START (Strategic Timing of Antiretroviral Treatment) studies. The SMART study showed that consistent use of ART in individuals with CD4^+^ cell counts below 350/µl resulted in a decrease in AIDS-related adverse events and in CVD events (Nsagha, Clement, *et al*., 2015).

For those deferring or interrupting treatment, there was a 70% increased hazard of CVD events, suggesting the need for continuous ART in preventing HIV-associated chronic inflammation and mitigating CVD risk (Vasan *et al*., 2016). In the START study, a 40% reduction in AIDS-related events was observed when ART was given immediately; however, this did not prevent CVD events. Although both SMART and START support the notion that ART on the basis of stricter CD4^+^thresholds will likely result in decreased CVD rates, ART is not sufficient to prevent CVD risk in HIV^+^ patients (Sniderman *et al*., 2011). Data from the VACS (Veterans Aging Cohort Study) cohort showed that HIV patients were at a higher risk of acute MI (hazard ratio, 1.48), even after adjustment for Framingham risk factors, comorbid conditions, and drug use. Thus, non-ART interventions are needed to decrease CVD risk among HIV^+^ populations and to improve immune function (Kiyotaka Hao, 2012).

HIV^+^ individuals known as “elite controllers,” that is, those who have undetectable plasma viral loads without ART(Feeney & Mallon, 2011), have increased coronary atherosclerosis and high immune activation, including elevated plasma soluble CD163 (sCD163). In another cohort, HIV^+^ elite controllers had a higher median carotid intima-media thickness (CIMT) than that observed in uninfected subjects, even after adjustment for traditional cardiovascular risk factors. These studies reinforce the role of inflammation, and not ART or virus, as the key mediator of HIV-associated CVD. Therefore, HIV-infected patients are at a higher risk of developing cardiovascular diseases compared to the general population (Vasan *et al*., 2016).

### 2.6. LABORATORY DIAGNOSIS

Before testing CVD, different factors are considered which includes; age, diet, family history of heart disease, physical activity, and blood pressure.

#### Lipid panel determination

This is done to determine the amount of lipids in the blood using manual or an automated machine like spectrophotometer and COBAS.

#### CRP

The elevation of level CRP in blood is associated with risks of heart disease (Yeh *et al*., 2003).

Some tests are done to determine the proteins that is released when muscle cells are damaged what we call cardiac biomarkers

#### Troponin

This test is done to indicate the degree of damage to the heart (KyungCP et al 2017).

#### Creatine Kinase-MB

This test is done to detect and monitor heart attacks during this case, CK is elevated. but nowadays; it has been replaced by troponin. In case of heart attacks CK is elevated (Mels *et al*., 2016).

Most cardiovascular disease are diagnosed based on history and physical examination (inspection, auscultion and palpation) (Abourbin s et al 2007).

Different methods are used to diagnose CVD such as:

**Electrocardiography** is specific in diagnosis of rhythm disturbances

**Echocardiography** is excellent to assess the severity of valvular regurgitations and stenotic lesions and to evaluate chamber enlargement and quantify systolic and diastolic myocardial function

**Lipid profile** determination by calculating the amount of lipid in the blood an automated machine

#### Thoracic radiographs

Are best to diagnose lungs for evidence of pulmonary hypertension.

### 2.7. TREATMENT OF RISK OF CARDIOVASCULAR DISEASES IN HIV/AIDS RECEIVING ART

#### Use of pravastatin or atorvastatin

They are very safely to be used in ART patients because of ability to reduce LDL-c and triglycerides in men and increase HDL-c in the blood. Both Pravastatin and atorvastatin: it includes to group of drugs known as “Statin”. (Schambelan *et al*., 2002).

#### Rosuvastatin

This is anti-inflammatory treatment that reduces mortality and venous thrombosis in people who have high levels of C-reactive protein (RidkerPM *et al*., 2008).

Other different ways can be used to treat the metabolic lipid changes such as:

#### Life style changes

This is concerned by avoiding tobacco in all means and choosing foods that are highly heart healthy „omega 3 and monosaturated fats and large amounts of dietary fiber. Regular physical exercises are also required.

#### Fibrates

For those patients who dot take Statin, doctors prescribe fibrates. This is used mostly to reduce the production of cholesterol (Abourbins *et al*., 2007).

## CHAPTER III: METHODOLOGY

This chapter describes the study area, study design, study population, sample size, sampling strategy, data collection methods and procedure, data analysis, problem and limitation and ethical consideration.

### 3.1. STUDY AREA

This study was conducted at University Teaching Hospital of Butare (CHUB) which is located in southern province, Huye district, Ngoma sector near university of Rwanda Huye campus and it was about 120 Km from Kigali city. The study has been carried out in clinical chemistry laboratory services where the lipid profile was determined.

### 3.2. STUDY DESIGN

Cross sectional was conducted during from January to February 2021 to assess the risks of cardiovascular diseases among HIV/AIDS infected individuals receiving antiretroviral therapy at University Teaching Hospital of Butare (CHUB) during 2021.

### 3.3. STUDY POPULATION

The study population were HIV/AIDS patients receiving ART at CHUB involving HIV/AIDAS positive cases and negative cases as control group.

### 3.4. SAMPLE SIZE

The sample size was calculated using the fisher formula**: N=T^2^(1-P) P/D^2^** (Ajay and Micah, 2014).

Where:

N: was the sample size required at 95% confidence interval level. T: was the coefficient interval at 95% which is 1.96.

P: this represents the prevalence of cardiovascular diseases in the population. D: was the margin error in the population which is 5%.

The prevalence of cardiovascular diseases in Rwanda was 15.3% (Nahimana *et al*., 2017) Therefore, with refer to this formula, **N=T^2^(1-P) P/D^2,^ N**=1.96^2^ [1-0.15.3]15.3/0.0025 Hence, the study recruited **199** participants as sample size.

### 3.5. INCLUSIVE CRITERIA

The study has included HIV/AIDS patients receiving ART at CHUB.

### 3.6. EXCLUSIVE CRITERIA

The study has excluded the participants with serological evidence of hepatitis B/C infection, abnormal thyroid hormone, pregnancy, hypertension, kidney disease, diabetes mellitus, alcoholism and obesity.

### 3.7. SAMPLING STRATEGY

Systematic sampling was used to recruit HIV/AIDS patients receiving ART at CHUB where a blood sample has been collected from these patients and processed in clinical chemistry laboratory for the lipid profile analysis. Participants‟ identifications were recorded in log book and they were given code of identification in order to maintain their confidentiality.

### 3.8. METHODS AND PROCEDURES

#### 3.8.1. Sample collection

In our study blood sample (serum) has been collected and analysed using the following materials:

- Dry tubes
- Gloves
- Tourniquets
- Sterile needles
- Vacutainer holder
- Alcoholic pads
- Waste bin
- Spectrophotometer machines (Humalyzer 3500)
- Test tube holders
- Plain tubes
- Cypress lipid profile kits (TC, TG, HDL and LDL)
- Distilled water
- Pipettes
- Tips
- Watch
- Papers
- Calculator
- centrifuge

#### 3.8.2. Procedures of the test

After collecting 199 blood samples, were examined in clinical chemistry laboratory under spectrophotometric humalyzer machine by

Using the following procedures:

##### 3.8.2.1. Procedure for cholesterol

- wavelength: 505nm/green
- temperature: 37°c/ at room temperature
- light path: 1cm Pipetting into clean dry test tube labelled as blank (B), standard (S) and test (T)
- working reagent was ready to be used, and 1000ul was added to blank, standard, and test tubes each.
- 10ul of distilled water was added to the blank labelled test tube
- 10ul of cholesterol standard was added to standard labelled test tube
- 10ul of serum sample was added also to the sample labelled test tube

(**Cholesterol cypress kit in accordance to manufacturer’s instructions)**

Solution has been mixed well and incubated at 37°c at room temperature for 15 minutes.

The absorbance of the standard and test against blank has been measured within 60 minutes.

#### Calculation

standard concentration=200mg/dl

Cholesterol in mg/dl= Abs T/Abs S * (200mg/dl)

##### 3.8.2.2. Procedure for triglyceride

- wavelength: 505nm/green
- temperature: 37°c/ at room temperature
- light path: 1cm Pipetting into clean dry test tube labelled as blank (B), standard (S) and test (T)
- working reagent was ready to be used, and 1000ul was added to blank, standard, and test tubes each.
- 10ul of distilled water was added to the blank labelled test tube
- 10ul of triglycerides standard was added to standard labelled test tube
- 10ul of serum sample was added also to the sample labelled test tube

**(Triglyceride cypress kit in accordance to manufacturer’s instructions)** Solution was mixed well and incubated at room temperature for 15 minutes.

The absorbance of the standard and test against blank has been measured within 60 minutes.

#### Calculation

Standard concentration=200mg/dl

Triglyceride in mg/dl= Abs T/Abs S *(200mg/dl)

##### 3.8.2.3. HDL cholesterol procedures

- wavelength: 505nm/green
- temperature: 37°c/ at room temperature
- light path: 1cm Pipetting into clean dry test tube labelled as blank (B), standard (S) and test (T)
- working reagent was ready to be used, and 1000ul was added to blank, standard, and test tubes each.
- 10ul of distilled water was added to the blank labelled test tube
- 10ul of HDL standard was added to standard labelled test tube
- 10ul of serum sample was added also to the sample labelled test tube

(HDL-**cypress kit in accordance to manufacturer’s instructions**) Solution was mixed well and incubated at 37°c for 5 minutes.

The absorbance of the standard and test against blank within has been measured 60 minutes.

#### Calculation

standard concentration=50mg/dl

HDL-Cholesterol in mg/dl= Abs T/Abs S *50mg/dl

##### 3.8.2.4. LDL-c procedures

- wavelength: 505nm/green
- temperature: 37°c/ at room temperature
- light path: 1cm
- Pipetting into clean dry test tube labelled as blank (B), standard (S) and test (T)working reagent was ready to be used, and 1000ul was added to blank, standard, and test tubes each.
- 10ul of distilled water was added to the blank labelled test tube
- 10ul of LDL-standard was added to standard labelled test tube
- 10ul of serum sample was added also to the sample labelled test tube

(LDL-**Cyprus kit in accordance to manufacturer’s instructions**) Solution was mixed well and incubated at 37°c for 5 minutes.

The absorbance of the standard and test against blank within has been measured 60 minutes.

#### Calculation

LDL-Cholesterol in mg/dl= Abs T/Abs S *50mg/dl

### 3.9. DATA ANALYSIS

The obtained data have been analysed using *Statistical Package for the Social Sciences*(SPSS) based on the result of lipid profile obtained and ARV drug taken by the participant. A P-value of ≤0.05 was considered statistically significant for any kind of comparison. The collected data was recorded by making use of Microsoft excel. Each patient‟s information regarding their age, sex, lipoprotein parameters including TC, HDL, LDL and triglycerides was recorded.

### 3.10. ETHICAL CONSIDERATION

The data were collected after getting Ethical clearance from University of Rwanda, College of Medicine and Health Sciences and approval from CHUB. All the participants voluntarily signed the informed consent after being explained the purpose of the study. The confidentiality was insured by using the code number and was only accessed by authorized personnel.

### 3.11. PROBLEMS AND LIMITATIONS

The findings of this study may not be generalizable to all age categories and all racial groups, as the majority of the sample were mainly southern province and the small sample size. Due to the COVID-19 Pandemic, Participants from different districts were not able to come to CHUB due to the movement restriction; hence they were recruited in this study. The delay to obtain the ethical approval at the hospital also impeded

## CHAPTER IV: RESULTS

### 4.1 DEMOGRAPHIC INFORMATION OF THE STUDY PARTICIPANTS

A total of 150 HIV/AIDS participants on ART were recruited in the study, 51 (34%) were males while 99 (66%) were females. The participants were on treatment for more than one year at the time of the study and they were between 29 and 75 years old with a mean age of 49.4 years. 108 (72%) participants were on first line medication whose 38 participants were male and 70 participants were females. 39 (26%) participants were on second line medication whose 10 participants were male and 29 participants were females. Among 49 (24.7%) participants serving as control group, 17 (34.7%) were males and 32 (65.3%) were females (Table 1).

**Table 1:** Participants characteristics: gender distribution and number of participants on ARV.

|  |  | ART line of treatment |  |  | Total n (%) |
| --- | --- | --- | --- | --- | --- |
|  |  | 1<sup>st</sup> line | 2<sup>nd</sup> line | 3<sup>rd</sup> line |  |
|  |  | n (%) | n (%) | n (%) |  |
| SEX | MALE | 38 (74.5) | 10 (19.6) | 3 (5.9) | 51 (34) |
|  | FEMALE | 70 (70.7) | 29 (29.3) | 0 (0.0) | 99 (66) |
|  | **TOTAL** | 108 (72.0) | 39 (26.0) | 3 (2.0) | 150 (100) |

### 4.2. LIPID PROFILE AMONG PEOPLE LIVING WITH HIV RECEIVING ART AT CHUB

#### 4.2.1. Triglycerides level

Among 150 participants who were receiving ART treatment regimens, 63(42.0) participants had a normal range of triglyceride and n=39(26.0) were borderline high levels whereas a total of 48(32%) participants had hypertriglyceridemia (Table 2).

**Table 2:** lipid profile distribution among participant‟s subjects receiving ART.

| Variables | ART regime |  |  | Total |
| --- | --- | --- | --- | --- |
|  | 1<sup>st</sup> line<br>n (%) | 2<sup>nd</sup>line<br>n (%) | 3<sup>rd</sup> line<br>n (%) | All lines<br>n(%) |
| **Triglyceride level (mg/dl)** |  |  |  |  |
| ≤150 : Normal range | 46 (42.6) | 15(38.5) | 2(66.7) | 63(42.0) |
| 151-199: Borderline high | 26 (24.1) | 12(30.8) | 1(33.3) | 39(26.0) |
| 200- 499 : Hypertrigylcemia | 36 (33.3) | 12 (30.8) | 0(0.) | 48(32) |
| Total: | 108 (100) | 39(100) | 3(100) | 150(100) |
| **LDL level (mg/dl)** |  |  |  |  |
| ≤130 : Normal range | 92 (85.2) | 33(84.6) | 2(66.7) | 127(84.6) |
| 131-159: Borderline high | 9 (8.3) | 2(5.1) | 0 (0.0) | 11(7.3) |
| 160 Above: Hypercholestemia | 7 (6.5) | 4 (10.3) | 1(33.3) | 12(8.0) |
| Total: | 108 (100) | 39(100) | 3(100) | 150(100) |
| **HDL level (mg/dl)** |  |  |  |  |
| ≤40 : Hypercholestemia | 23 (21.3) | 5(13.2) | 1(33.3) | 29(19.3) |
| 40-59: normal | 20 (18.5) | 8(18.4) | 1(33.3) | 29(19.3) |
| Above 60 : low risks | 65 (60.2) | 26 (68.4) | 1(33.3) | 92(61.3) |
| Total: | 108 (100) | 39(100) | 3(100) | 150(100) |
| **TC level( mg/dl)** |  |  |  |  |
| ≤200 : Normal range | 63 (58.3) | 28(71.8) | 0(0.0) | 91(60.6) |
| 201-239: Borderline high | 29 (26.9) | 6(15.4) | 1(33.3) | 36(24.0) |
| 240 Above: Hypercholestemia | 16 (14.8) | 5 (12.8) | 2(66.7) | 23(15.3) |
| Total: | 108 (100) | 39(100) | 3(100) | 150(100) |

#### 4.2.2. High density lipoprotein cholesterol level

In all 150 participants who were receiving ART treatment, regimens, n=92(61.3%) were normal and n=29 (19.3%) were classified to have low risks for developing CVD whereas n= 29 (19.3%) of all medication levels were hypercholestemia and the majority were receiving first line medication (Table 2).

#### 4.2.3. Low density lipoprotein cholesterol

Upon all 150 participants who were receiving ART treatment regimens, 127(87.6) participants had the normal range of LDL-C and n=11(7.30) were borderline high for LDL-C measurements whereas n= 12(8%) of all medication levels were hypercholestemia (Table 2).

#### 4.2.4 Total cholesterol level

Among 150 participants who were receiving ART treatment regimens, 91(60.6) participants had normal range of Total Cholesterol and 36 (24) participants were borderline high for total cholesterol measurements whereas a total of 23 (15.3) participants had hypercholestemia of all medication levels (Table 2).

### 4.3. ASSOCIATION BETWEEN LIPID PROFILE AND ART TREATMENT REGIMENS USED IN HIV/AIDS

Lipid profile among participants under ART treatment were compared with the control group representing individuals who were HIV negative and obviously who did not take the ART. For triglycerides level measurement, in contrary to participants under ART, there was no any case of high triglycerides level in control group and almost all participants [44 (89.8)] within this group had the normal range of the triglyceride. (Table 3) The P value at this level was less than 0.046 (P value <0.046), meaning that there is significance increase of triglycerides among the participants who were receiving ART compared to the control group (Table 3).

**Table 3:** Association between lipid profile and ART treatment regimens used in HIV/AIDS patient.

|  | ART regime |  |  | Total<br>All lines<br>n(%) | P-value<br>For<br>ART | Control<br>Group<br>n(%) | P-value<br>for<br>Contro<br>l group |
| --- | --- | --- | --- | --- | --- | --- | --- |
| Variables | 1<sup>st</sup> line<br>n (%) | 2<sup>nd</sup>line<br>n (%) | 3<sup>rd</sup> line<br>n (%) |  |  |  |  |
| **Triglyceride level (mg/dl)** |  |  |  |  |  |  |  |
| ≤150 : Normal range | 46 (42.6) | 15(38.5) | 2(66.7) | 63(42.0) |  | 44 (89.8) |  |
| 151-199: Borderline high | 26 (24.1) | 12(30.8) | 1(33.3) | 39(26.0) | **0.046** | 5(10.2) | **0.11** |
| 200-499 : Hypertrigylcemia | 36 (33.3) | 12 (30.8) | 0(0.) | 48(32) |  | 0 |  |
| Total: | 108 (100) | 39(100) | 3(100) | 150(100) |  | 49(100) |  |
| **LDL level (mg/dl)** |  |  |  |  |  |  |  |
| ≤130 : Normal range | 92 (85.2) | 33(84.6) | 2(66.7) | 127(84.6) |  | 43 (84.7) |  |
| 131-159: Borderline high | 9 (8.3) | 2(5.1) | 0 (0.0) | 11(7.3) | **0.039** | 5(12.2) | **0.13** |
| 160 Above: Hypercholestemia | 7 (6.5) | 4 (10.3) | 1(33.3) | 12(8.0) |  | 1(3.1) |  |
| Total: | 108 (100) | 39(100) | 3(100) | 150(100) |  | 49(100) |  |
| **HDL level (mg/dl)** |  |  |  |  |  |  |  |
| ≤40 : high risks | 23 (21.3) | 5(13.2) | 1(33.3) | 29(19.3) |  | 4 (8.2) |  |
| 40-59: normal | 20 (18.5) | 8(18.4) | 1(33.3) | 29(19.3) | **0.021** | 21(42.9) | **0.099** |
| Above 60 : low risks | 65 (60.2) | 26 (68.4) | 1(33.3) | 92(61.3) |  | 24(49.8) |  |
| Total: | 108 (100) | 39(100) | 3(100) | 150(100) |  | 49(100) |  |
| **TC level( mg/dl)** |  |  |  |  |  |  |  |
| ≤200 : Normal range | 63 (58.3) | 28(71.8) | 0(0.0) | 91(60.6) |  | 44 (86.7) |  |
| 201-239: Borderline high | 29 (26.9) | 6(15.4) | 1(33.3) | 36(24.0) | **0.045** | 3(7.2) | **0.16** |
| 240 Above: Hypercholestemia | 16 (14.8) | 5 (12.8) | 2(66.7) | 23(15.3) |  | 2(6.1) |  |
| Total: | 108 (100) | 39(100) | 3(100) | 91(60.6) |  | 49(100) |  |

For low LDL-C, a great number [12(8.0%)] of people who were on ART treatment, had a high level of LDL-C compared to control group [(1(3.1%)]. The P value at this level was less than 0.039(P value <0.039), meaning that there is clinical significance to the increase of LDL among these participants who were receiving ART compared to the control group with P value (0.16) (Table 3). There is also great number of participants 29(19.3) who had a low level of HDL-C compared to the control group [4 (8.2%)]. The P value at this level was less than 0.021 (P value <0.021), meaning that there is clinical significance to the increase of HDL among these participants who were receiving ART compared to the control group whose P value (0.16) (Table 3).

For Total cholesterol, among the individual who were under ART 23 (15.3%) participants, had a high level of Total cholesterol which is a high number compared to control group which had only 2 case representing 6.1%. The P value at this level was less than 0.045 (P value <0.045), meaning that there is clinical significance to the increase of total cholesterol among these participants who were receiving ART compared to the control group (Table 3).

## CHAPTER V: DISCUSSION

Lipid metabolism abnormalities have been increasingly recognised among HIV-infected patients treated with HAART. Dyslipidaemia is therefore very common among HIV positive people that are on (HAART) and the association of having this lipid changes due to taking ART varies between 8% to 32%. During our study, we have found that 15(8%) participants receiving ART (all treatment lines) were having high LDL in correlation with taking ART regimens as the risks of developing CVDs, 29(19.3%) participants who were receiving ART at all treatment lines have been found to have high HDL(hypercholestemia) in response of taking these medications, 23(15.3%) hypercholestemia(high TC) cases have been found in our study as well as 48(32%) Hypertrigylcemia (high TG) cases have been identified during our study in relation of receiving ART regimens.

A correspondence of having high LDL-c in our study was in disagreement (lower than) in that one of reported in the study carried out in urban Cameroon which was (43.5 %). The increase of LDL level among urban participants in Cameroons, were mainly due urban life style including feeding habit as urban population tends to eat more fat food especially from industries which contributed to abruptly increase of that LDL among Cameroonian population (Bekolo *et al*., 2014). Within our study the participants were coming from rural areas around CHUB which made this LDL derangements due to ART to be lower than that of urban Cameroonians. High LDL due to receiving ART reported from India (16.1%) (WHO,2005) was also in disagreement (twice) in that of our study, the participants were receiving different medications and most of their study population were mainly receiving ART for a period above to 8 years, as in our study most of the participant‟s period of receiving ART regimens was ranging from 3 to 5 years.

We have found that the participants with high TC due to ART was (15.3%). This Correlation rate was higher than and in disagreement as that of the study reported from a West African which was 4.5% (Ogundahunsi, O etal, 2010). Based on HDL-c cut off value, high HDL-c level (hypercholestemia) in our treatment group was (15.3%). However, it is less than and in disagreement of that reported from rural South India which was 24.8% (Bernal E et al,2013) as they study participants were mainly coming from different areas and their living style was different as in our study population were coming from the same area, hence makes this rate to be different.

We have found that, the participants with raised TG due to receiving ART in our study was 32%. This is not in line with that one reported from Kenya, and India. The variation of TG due to receiving ART in these two studies were respectively, 43.5%, and 42.1%. According to our study observation TG findings, were in beyond of their findings as during their study, they did not exclude participants with obesity and alcoholic participants were permitted to intervene in their study as these factors could increase the rate of having high TG as in our study we have excluded these participants based on exclusion criteria, hence we did not match our findings (Karari *et al*., 2014).

However, there are suggestions that the magnitude of lipid profile derangements induced by ART could show variation with duration of treatment, across populations and setting (Betyoumin *et al*., 2011).

## CHAPTER VI: CONCLUSIONS AND RECOMMENDATIONS

This chapter concludes the research report and help us to take conclusion from data collected during research in relation to the aims of the study and recommendations in regard the need to test HIV positive patients for CVD.

### 6.1. CONCLUSIONS

The findings of this research indicate that there is significance increase of lipid profile among HIV patients that are receiving HAART treatment regimens.

This study found that there is high level of lipid profile due to receiving ART.we have found that 15(8%) participants receiving ART (all treatment lines) were having high LDL in correlation with taking ART regimens as the risks of developing CVDs, 29(19.3%) participants who were receiving ART at all treatment lines have been found to have high HDL(hypercholestemia) in response of taking these medications,23(15.3%) hypercholestemia(high TC) cases have been found in our study as well as 48(32%) Hypertrigylcemia (high TG) cases have been recognized during our study in relation of receiving ART regimens. Principally these participants were mainly receiving first line of medications. Therefore, this study should be conducted within the other health institutions in HIV/AIDS management at different districts and other Referral hospitals to have this association in hand for more concerned population.

The findings of this study suggest that there is significance change in lipid profile among HIV infected individuals who are receiving ART treatment regimens.

### 6.2. RECOMMENDATION

With refer to the findings, to assess the risks of cardiovascular among HIV/AIDS patients receiving ART, we acclaim the other future researcher to this association to perform extra tests for CVD management such as troponin, Creatine Kinase-MB, CRP as well as electrolytes measurements additionally to lipid profile testing. The information generated from the study have to support the Ministry of Health as well as different referral hospitals and district hospitals in policies development regarding the prognosis and management of cardiovascular diseases in HIV/AIDS patients receiving antiretroviral therapy. Therefore, detection of lipid profile in PLWH may be added to the routine biological assessment to determine the risks of CVD for early intervention and prevention of the disease.

## Data Availability

All data produced in the present work are contained in the manuscript

## Notes

### Competing Interest Statement

The authors have declared no competing interest.

### Funding Statement

This study did not receive any funding

### Author Declarations

The data were collected after getting Ethical clearance from University of Rwanda, College of Medicine and Health Sciences and approval from CHUB.

